# A Retrospective Analysis of Serious Adverse Events and Deaths in US-Based Lifestyle Clinical Trials for Cognitive Health

**DOI:** 10.1101/2023.09.27.23296243

**Authors:** Mickeal N. Key, Ashley R. Shaw, Kirk I. Erickson, Jeffrey M. Burns, Eric D. Vidoni

## Abstract

This retrospective analysis assessed the serious adverse events and deaths reported in lifestyle clinical trials designed to enhance cognitive health in older adults living in the United States. Data was collected from studies conducted between January 1, 2000, and July 19, 2023, using the ClinicalTrials.gov application programming interface. Our query revealed that 76% of these studies did not report trial results. The remaining studies with reported results were categorized under one of four intervention types: Cognitive/Behavioral, Exercise/Movement, Diet/Supplement, and Multi-modal. When all trial types are considered together, the results indicate that lifestyle clinical trials are safe, with no significant increase in relative risk of experiencing an SAE in an intervention group over a control group. And although the increase in relative risk of death in an intervention group over a control group was significant at 28% (X^2^ (1, N = 36), p < 0.00688), the probability of death was not higher than the U.S. mortality rates by age. When assessing the data using intervention type, Diet/Supplement trials and Multi-modal trials both had an increase in relative risk of experiencing an SAE in the intervention over the control group, with Diet/Supplement trials at 16% (X^2^ (1, N = 2), p < 0.0263) and Multi-modal trials at 365% (X^2^ (1, N = 5), p < 0.000213). The Diet/Supplement trials also had an increased risk of death at 67% (X^2^ (1, N = 2), p < 0.000197). These results should be taken with careful consideration. Due to such a low reporting rate, the 36 studies included in this analysis do not accurately represent the majority of lifestyle clinical trials conducted in the U.S. This study is valuable in that it highlights the importance of reporting clinical trial results, which will improve transparency in trial results and allow for more accurate assessments of safety in the growing field of cognitive aging and lifestyle interventions for older adults.

## Introduction

Over the last two decades, many well-controlled clinical trials have improved our understanding of the potential for lifestyle modifications such as diet, exercise, meditation, and cognitive training to enhance cognitive health ^[**1**]^. A recent World Health Organization analysis estimated that up to 40% of dementia cases are attributable to modifiable factors, and approximately 10% of those are potentially related to physical health behaviors ^[**2, 3**]^.

Continued investigation of behavioral, non-pharmaceutical clinical trials for cognitive health warrants an assessment of their safety. Common familiarity with healthy behaviors such as walking or eating more vegetables is not equivalent to safe study conduct and should not be assumed to be safer than investigational medications. Conversely, it should not be assumed that lifestyle interventions for cognitive health, generally focused on older adults, pose an elevated risk for participants. Institutional review boards, oversight committees, and investigative teams all need reliable data to adequately assess the risk associated with these interventions.

The safety of a clinical trial is often measured in terms of the frequency, severity, and relatedness of adverse events. An adverse event (AE) is commonly defined as any physical or psychological sign, symptom, or disease experienced by a research participant during the study period. A serious adverse event (SAE) is commonly defined as any adverse event that results in death, is life-threatening, requires hospitalization, causes disability, or poses a significant hazard. A standard approach for assessing and reporting both AEs and SAEs is provided by the NIA Adverse Event and Serious Adverse Event Guidelines, Version 5.

The goal of this retrospective analysis was to evaluate the safety of reported lifestyle clinical trials for cognitive health in older adults aged 60 and older in the United States by assessing 1) serious adverse events (SAE) and 2) deaths in lifestyle clinical trials. Our hypothesis was that participants in intervention groups would experience a higher risk of SAE (including death) than participants in control groups. Due to the characteristics of the study population, older adults, we also hypothesized that interventions requiring exercise would experience a higher risk of SAE (including death) than any other lifestyle intervention type. Our intended outcome was to provide context for the current state of safety in lifestyle clinical trials, ideally to inform future trial design, implementation, and dissemination of trial results.

## Methods

Data on the frequency of SAE and deaths in lifestyle clinical trials was extracted from ClinicalTrials.gov using their application programming interface (API). We limited our search to registered clinical trials completed between January 1, 2000 and July 19, 2023. Other search criteria included 1) minimum age of 60 years for inclusion criteria, 2) at least one explicitly identified outcome of cognitive function, 3) randomized allocation, 4) intervention with a clearly identifiable control condition, and 5) a United States-based coordinating location.

The search string supplied to the API was constructed with consideration for prior systematic reviews ^[**1, 4-7**]^, and included:

### Intervention/Treatment

Physical Activity OR Exercise OR Sports OR Walking OR Biking OR Bicycling OR Running OR Fitness OR Yoga OR Strength OR Resistance OR Aerobic OR Endurance OR Flexibility OR Tai Chi OR Qi Gong OR Balance OR Diet OR Dietary OR Nutrition OR Nutritional OR Food OR Supplement OR Vitamin OR Multivitamin OR Cognitive OR Cognition OR Reading OR Games OR Gaming OR Mindfulness OR Meditation OR Sleep OR Dance

### Outcome measure

Cognition OR Memory OR Executive OR Thinking OR Cognitive OR Attention OR Processing

We defined lifestyle interventions as those designed to improve physical and/or cognitive health. This definition excludes interventions where a drug or device initiates the physical or cognitive change. Interventions using natural products or therapeutic compounds requiring a prescription were also excluded. Due to the broad nature of lifestyle interventions, it was necessary to categorize similar interventions into categories. For this review, two investigators (MNK, EDV) reached consensus on assignment of each study to one of four broad lifestyle intervention categories, referred to as “Intervention Type”. The “Cognitive/Behavioral” category included any intervention that involved cognitive training, brain games, sensory interventions, meditation, and assessment/screening interventions. The “Exercise/Movement” category included any intervention that involved exercise, yoga, dance, stretching/toning or similar. The “Diet/Supplement” category included any study that involved feeding, dietary changes, supplements, and bioactive natural products. Finally, the “Multi-modal” category included any intervention that made use of two or more intervention categories (e.g., an intervention that had participants undergo cognitive training and exercise could be considered Multi-modal).

For each included study, the number of individuals affected and at risk of experiencing an SAE, as well as the number of individuals who died during the study period, were evaluated. To provide additional context, risk and risk ratios were calculated for SAEs and deaths, both in studies overall and by intervention type. The risk of an event for intervention and control group was calculated as the number of participants affected divided by the number of participants at risk. The risk ratio was calculated as the risk of an event in the intervention divided by the risk of an event in the control group. The risk ratios are accompanied by 95% confidence intervals and a chi-square test to evaluate whether there was a statistically significant difference between intervention and control groups. The API query and statistical analyses were all performed using R Statistical Software^[**8**]^.

## Results

Of the 6,497 lifestyle intervention studies on ClinicalTrials.gov that met the review criteria, 6461 were excluded from analysis: 4953 did not have results posted; 885 were not lifestyle interventions as described in the previous section; 589 included individuals below age 60; 19 had no cognitive outcome listed; 13 were centralized outside the US; 2 did not include random allocation. Thirty-six (36) studies remained for analysis.

Of these 36 included studies, all reported SAEs and 30 studies (83%) reported deaths. The median study duration for all studies was 6 months, with the shortest intervention period lasting 2 hours (0.00274 months) and the longest intervention lasting 10 years (120 months). The percentage of participants experiencing a SAE in the included lifestyle clinical trials was 5.13%. The percentage of deaths during the study period for the included studies was 2.21%. Table 1 includes the number of studies in each of the four intervention types, the study duration, and documents the number of SAEs and deaths for each, as well as overall.

**Table 1.** Studies Reporting SAEs and SAE Counts by Intervention Type.

|  | **Overall** | **Cognitive/**<br>**Behavioral** | **Diet/**<br>**Supplements** | **Exercise/**<br>**Movement** | **Multi-Modal** |
| --- | --- | --- | --- | --- | --- |
| Number of Studies, n<sup>1</sup> | 36 | 18 | 2 | 11 | 5 |
| Median Length of Studies in<br>Months, n (range) | 6<br>(0.00274-120) | 6<br>(0.00274-120) | 18.2<br>(2.76-33.6) | 5.98<br>(0.23-18) | 8.49<br>(2.76-18) |
| Participants at Risk, n | 21,042 | 8,624 | 10,289 | 1,082 | 1,047 |
| **Serious Adverse Events** |  |  |  |  |  |
| Studies Reporting SAEs, n (%) | 36 (100%) | 18 (100%) | 2 (100%) | 11 (100%) | 5 (100%) |
| Participants Affected by SAE, n<sup>2</sup> | 1,079 | 182 | 765 | 78 | 54 |
| Number of SAE, n<sup>3</sup> | 1,111 | 83 | 942 | 32 | 54 |
| **Deaths** |  |  |  |  |  |
| Studies Reporting Deaths, n (%) | 29 (81%) | 14 (78%) | 2 (100%) | 9 (82%) | 4 (80%) |
| Number of Deaths, n | 466 | 256 | 201 | 3 | 6 |
| <sup>1</sup>Meeting inclusion criteria for analysis. |  |  |  |  |  |
| <sup>2</sup>Number of unique participants who were affected by at least one serious adverse event (SAE), not including death. |  |  |  |  |  |
| <sup>3</sup>Total Number of SAE, not including deaths. Number of SAE may be more than number of unique participants affected in cases where a participant had more than one SAE. However, we noted that two cognition intervention trials and one exercise intervention trial incorrectly underreported the total number of SAE vs the total number of participants affected resulting in the number of SAE being lower than the number of participants affected by SAE. |  |  |  |  |  |

The overall risk ratio for lifestyle interventions describes a 5.8% relative decrease in risk for experiencing an SAE in the intervention group compared to the control group (0.941 times as much probability). This decrease in risk was not statistically significant, as evidenced by a chi-square test, X^2^ (1, N = 36), p < 0.308. When SAE risk was assessed by intervention type, Diet/Supplement and Multi-modal studies were the only ones to have statistically significant risk ratios. Both had SAE risk ratios over 1, indicating an increased relative risk of experiencing an SAE in the intervention over control, with Diet/Supplement studies at an increased relative risk of 16% (X^2^ (1, N = 2), p < 0.0263) and Multi-modal studies at an increased relative risk of 365% (X^2^ (1, N = 5), p < 0.000213).

When considering overall deaths during the study duration of included trials, the risk ratio describes a 28% relative increase in risk of death in the intervention group than in the control group (1.28 times as much probability). This increase in risk was statistically significant (X^2^ (1, N = 36), p < 0.00688), and translates to a 2.98% probability of death in the intervention group and a 2.33% probability of death in the control group. This finding was likely driven by Diet/Supplement studies, which was the only intervention type to have a statistically significant risk ratio at 67% (X^2^ (1, N = 2), p < 0.000197). Risk and risk ratios, along with 95% confidence intervals and chi-square p-values across all studies, and separated by intervention type, are described in Table 2.

**Table 2.** Risk RaGo for Serious Adverse Events (SAEs) & Death by IntervenGon Type.

| Table 2. Risk Ratio for Serious Adverse Events (SAEs) & Death by Intervention Type |  |  |  |  |  |  |  |  |  |  |  |  |
| --- | --- | --- | --- | --- | --- | --- | --- | --- | --- | --- | --- | --- |
|  | SAEs |  |  |  |  |  | Deaths |  |  |  |  |  |
|  | Individuals w/ SAE | Individuals At Risk | Risk | Risk Ratio | Confidence Interval | Chi-Square *p*-value | Individuals Died | Individuals At Risk | Risk | Risk Ratio | Confidence Interval | Chi-Square *p*-value |
| **Cognitive/Behavioral** |  |  |  |  |  |  |  |  |  |  |  |  |
| Intervention | 99 | 5143 | 0.0192 | 0.807 | 0.605,1.08 | 0.145 | 131 | 2661 | 0.0469 | 0.993 | 0.782,1.26 | 0.958 |
| Control | 83 | 3481 | 0.0238 |  |  |  | 125 | 2522 | 0.0472 |  |  |  |
| **Diet/Supplement** |  |  |  |  |  |  |  |  |  |  |  |  |
| Intervention | 346 | 4262 | 0.0812 | 1.16 | 1.02,1.34 | 0.0263 | 109 | 4153 | 0.0255 | 1.67 | 1.27,2.21 | 0.000197 |
| Control | 419 | 6027 | 0.0695 |  |  |  | 92 | 5935 | 0.0152 |  |  |  |
| **Exercise/Movement** |  |  |  |  |  |  |  |  |  |  |  |  |
| Intervention | 39 | 592 | 0.0659 | 0.810 | 0.529,1.24 | 0.335 | 1 | 506 | 0.0019 | 0.441 | 0.0402,4.85 | 0.492 |
| Control | 39 | 480 | 0.0813 |  |  |  | 2 | 447 | 0.0044 |  |  |  |
| **Multi-modal** |  |  |  |  |  |  |  |  |  |  |  |  |
| Intervention | 49 | 710 | 0.0690 | 4.65 | 1.87,11.6 | 0.000213 | 5 | 692 | 0.0072 | 2.34 | 0.275,20.0 | 0.420 |
| Control | 5 | 337 | 0.0148 |  |  |  | 1 | 325 | 0.0030 |  |  |  |
| **Overall** |  |  |  |  |  |  |  |  |  |  |  |  |
| Intervention | 533 | 10,707 | 0.0498 | 0.941 | 0.838,1.06 | 0.308 | 246 | 8006 | 0.0298 | 1.28 | 1.07,1.53 | 0.00688 |
| Control | 546 | 10,325 | 0.0520 |  |  |  | 220 | 9226 | 0.0232 |  |  |  |

## Discussion

Our retrospective analysis of reported serious adverse events (SAEs) and deaths across more than 20 years of reported clinical trial results suggests a high degree of safety within lifestyle clinical trials related to cognitive health in older adults. The increased use of Consolidated Standards of Reporting Trials (CONSORT) guidelines ^[**9**]^ has likely improved safety reporting in the last decade, especially with the extended guidelines for safety published in 2004 ^[**10**]^. Our analysis also revealed significant gaps and irregularities in reporting of SAEs (including deaths) by the field, with 76% of completed studies in our ClinicalTrials.gov query not posting results.

The overall probability of an enrollee experiencing at least one SAE was about 5%, and was consistent between intervention and control conditions. The comparable risk between SAEs occurring in the intervention group versus the control group overall is encouraging but should be interpreted with caution. We were unable to evaluate the relatedness of SAE to intervention, a common classifier in safety reporting. Rather we presumed that if lifestyle interventions conveyed significant additional risk, that risk would be identifiable in the aggregate risk ratio. A higher risk ratio of intervention was noted for both the Diet/Supplement and Multi-modal interventions, though this must be considered carefully, as both categories had 2 and 5 studies, respectively, to evaluate.

While the overall risk of death for participants in an intervention group was statistically higher than those in a control group, the probability was not higher than the U.S. mortality rate. Between 1% and 5% probability of death in a study may appear concerning, until one considers that the probability of death in a given year for a 60-year-old in the US is between 0.7% and 1.3% and steadily increases with age. For a 75-year-old, the probability of death in a given year is 2.77% for women and 4.06% for men, climbing to 8.3% and 10.9% respectively by 85, a common upper age limit for trials ^[**11**]^.

The data also raise the potential of reporting biases inherent in the execution of trials. For example, the Multi-modal and multi-armed Diet/Supplement studies may create more contact points for participants to report SAE to study staff. Conversely, preconceived expectations about the safety of exercise for older adults may result in closer monitoring of participants, and an artificially lower probability of SAE in Exercise interventions, due to earlier intervention modification or termination at sub-SAE threshold events.

There are limitations to this retrospective analysis. First, not all studies reported results uniformly across our analysis period. We relied on reporting in ClinicalTrials.gov, which has only been required for all drug, biologic, and device clinical trials regulated by the U.S. Federal Drug Administration since 2017^[**12**]^. This requirement also applies to all clinical trials funded by the National Institutes of Health (NIH) and the Department of Veterans Affairs (DVA). Although the database supports semi-standardized reporting, we found that not all studies reported SAE and deaths. This inconsistency was recently noted in the updated CONSORT Harms 2022 guidance^[**13**]^. Broad uptake and universal implementation of these standardizing recommendations would improve our understanding of safety in our clinical research and in our interventions under investigation ^[**14**]^. Another factor in the number of studies included in our analysis is the fact that most lifestyle clinical trials would not fall under FDA regulation, which ultimately means that our sample is almost exclusively NIH and DVA-funded trials. While trials with non-federal and industry funding may register in ClinicalTrials.gov, they are not obligated to comply with established policies on the submission of study results.

Regardless of the reason, insufficient reporting of clinical trial results undermines transparency in the scientific process, our commitment to the responsible conduct of research, hinders trust building with the general public, and impedes the ability to monitor trends in lifestyle clinical trials that could reveal equity issues not readily apparent in other data such as intervention effectiveness or adherence. Underreporting is a source of bias for the field and a disservice to the participants who accepted risk for medical advancement, and the public who often funded the work.

Despite these limitations, our analysis has important implications for the growing field of cognitive aging and lifestyle clinical trials. Cognitive health studies have been a major focus of National Institute of Health funding in recent years. NIH requires a high degree of oversight and reporting, including data and safety monitoring committees, trial registration, and regular reporting. Additionally, cognitive health studies for people without dementia typically focus on older adults, for whom there is often greater concern regarding lifestyle modifications. Understanding the safety of these studies is and will continue to be paramount as the field grows. Though our estimates do not inform population-wide estimates of death and SAE, of which the literature is scant, the results presented are similar to the reported incidence of sudden death with physical activity in a general population of all ages ^[**5**]^. As investigators, we are regularly questioned by participants, Data and Safety Monitoring Committees, Institutional Review Boards, and others, as to the safety of lifestyle interventions, without sufficient data. We strongly recommend all clinical trials fully report study results to clinicalTrials.gov and adopt the CONSORT Harms 2022 guidance for future reporting, regardless of funding source. We expect these steps will lead to greater trustworthiness and rigor in cognitive health trials going forward.

## Data Availability

All data produced are available online at clinicaltrials.gov

## Declaration of financial/other relationships

The authors have no financial or other relationships to declare related to this manuscript.

## Author contributions

Conceptualization: EDV; Methodology: MNK and EDV; Writing, Review, & Editing: MNK, ARS, KIE, JMB, and EDV; Supervision & Project Administration: JMB and EDV. All authors have read and agreed to the published version of the manuscript.

## Acknowledgments

No assistance in the preparation of this article is to be declared

